# Physical activity and tobacco smoking in the German adult population

**DOI:** 10.1101/2024.08.19.24312219

**Authors:** Zeynep Acar, Sarah Jackson, Stephanie Klosterhalfen, Daniel Kotz

## Abstract

**Background:** Physical inactivity and tobacco smoking remain leading causes of morbidity and mortality worldwide. In Germany, smoking prevalence is high at around 30% and only 45% achieve the World Health Organization recommendation for physical activity (PA). Understanding how smoking and physical inactivity co-occur can inform interventions targeting these behaviours.

**Methods:** We analysed data from 4,073 adults (≥18 years) participating in a national household survey between April and July 2022. We tested the association between PA level (not=reference/low/medium/very active) and smoking status (never=reference/ex/current smoker). Among current smokers, we tested associations between PA level and cigarettes smoked per day; time spent with, and strength of, urges to smoke; and motivation to stop smoking.

**Results:** Overall, 29.9% (95%CI=28.5;31.4) reported no leisure time PA; among current smokers it was 39.8% (95% CI=37.3;42.4). Higher PA levels were associated with lower odds of being a current vs. never smoker (OR=0.74, 95%CI=0.69;0.79). Among current smokers, higher PA levels were associated with smoking fewer cigarettes per day (β=-0.98,95%CI=- 1.39;-0.56), weaker urges to smoke (OR=0.81,95%CI=0.74;0.89), and higher motivation to stop smoking (OR=1.13,95%CI=1.02;1.24). Time spent with urges to smoke showed a small overlap in the upper CI (OR=0.93,95%CI=0.85;1.02).

**Conclusion:** People who are more physically active are less likely to smoke. Current smokers with higher PA levels smoke less, are less dependent, and more motivated to quit. Further research is required to determine whether these associations are causal and, if so, whether interventions to increase PA could help people to quit smoking.

**What is already known on this topic:** Smokers are more likely to be physically inactive than non-smokers. The more cigarettes they smoke, the greater their risk of being physically inactive. Therefore, people who are physically inactive and smoke cigarettes have an increased risk of chronic diseases and premature death.

**What this study adds:** Our study adds detailed data on the relationships between physical activity (PA) levels and smoking habits in the German adult population. To build on existing studies that have explored the link between PA levels and either smoking status or the amount of cigarettes smoked per day, we also examined the associations between PA levels and both nicotine dependence and the motivation to quit smoking.

**How this study might affect research, practice or policy:** These results indicate a clear need to increase PA levels in the population and reduce smoking prevalence. Understanding the relationships between PA levels and smoking habits can aid in developing prevention strategies and targeting specific groups, such as current smokers. Additionally, this knowledge can inform the design of effective public health campaigns on PA and smoking, highlighting the association between these behaviours. More research is needed to determine whether associations are causal.

## INTRODUCTION

Tobacco smoking and physical inactivity are leading causes of morbidity and mortality worldwide [1-3]. Globally, tobacco use is the second leading risk factor for mortality with 5.1 million deaths and physical inactivity the fourth leading risk factor with 3.2 million deaths per year, and are responsible for chronic diseases such as heart and lung disease [4]. People who are highly physically active, but smoke tobacco, are still at increased risk of these diseases [3]. Therefore, smoking cessation and increasing physical activity (PA) levels should be public health goals for disease prevention and increased longevity [5, 6].

In Germany, the prevalence of smoking has remained high for years and is currently over 30% [7]. In addition, a 2017 study showed that less than half (45%) of the population met the World Health Organization (WHO) recommended minimum levels of PA [8]. This includes 150 to 300 minutes of moderate aerobic activity or 75 to 150 minutes of vigorous aerobic activity per week, plus strength training twice a week [9].

Some representative studies have reported associations between PA during leisure time and smoking status [3, 10, 11]. For example, in a survey from Switzerland with more than 18,000 adults, smokers were more likely to report low leisure time PA than non-smokers [10]. This study also provided evidence of a dose-response relationship, whereby smokers with higher cigarette consumption had a higher risk of low PA [10]. A study of over 59,000 adults from Norway also reported that smokers had a lower prevalence of PA than non-smokers, but only vigorous and not moderate PA was assessed [11]. However, these studies did not examine associations between different smoking behaviours and PA levels.

In smoking cessation, the motivation to stop and the level of nicotine dependence (which can be measured, among other things, by nicotine craving [12], defined here as the urges to smoke) are important factors in attempting to quit and in the success of such an attempt [13, 14]. Acute bouts of exercise have been shown to be associated with increased motivation to stop smoking, higher rates of abstinence in the last 24 hours, and reduced urges to smoke [14-17]. This suggests that PA interventions may have an effect on the motivation to stop smoking and the urges to smoke, at least in the short term. A recent review recommended that PA should be included in smoking cessation programmes [18]. However, whether current cigarette smokers with higher PA level in everyday life also show weaker urges to smoke and higher motivation to stop smoking has not yet been investigated through population studies. Knowledge of PA and smoking behaviour in the population allows general conclusions to be drawn without assuming laboratory conditions.

Therefore, we aim to expand the current evidence base and answer the following research questions:

1. What is the association between leisure time PA level (not=reference/low/medium/very active) and smoking status (never=reference/ex/current smoker) in the German adult population?
2. In the subgroup of current smokers, what is the association between leisure time PA level and (a) number of cigarettes smoked per day; (b) urges to smoke (as an indicator of nicotine dependence); and (c) motivation to stop smoking?

## METHODS

### Study design and setting

A study protocol was published on the Open Science Framework (https://osf.io/cmvk6). Data were used from the German Study on Tobacco Use (DEBRA study), a representative cross-sectional survey conducted on an ongoing basis, with questions primarily relating to the use of tobacco and alternative nicotine delivery systems in the German population (https://osf.io/4h92j/). Every two months, the sample is expanded by approximately 2,000 participants (= one survey wave). The computer-assisted, face-to-face interviews are conducted by an experienced market research institute. Respondents are selected using a dual frame design: an approximately 50:50 mix of multiple stratified, multistage random sampling and quota sampling (further information: https://osf.io/e2nqr/). Data on leisure time PA were collected in two DEBRA waves in 2022 from 28.04-29.05 (wave 36) and 23.06-24.07 (wave 37).

### Outcome measures

To measure PA, a single item was chosen for economic reasons. All participants were asked: “In a normal week, how much time in total do you spend on physical activity that increases your breathing rate? This may include sports, gymnastics, and brisk walking or bicycling for leisure or transportation. It does not include housework, gardening or physical activities that may be part of your job.” The respondents could indicate the answer in minutes or hours. The question had been adapted from Milton and colleagues [19, 20]. Following the authors, we based the item on leisure time PA. PA can be considered from different perspectives, such as leisure time PA, which is done by choice, or occupational PA, which is not done by choice and may not provide the same health benefits as leisure time PA [21-24]. As WHO recommendations for PA are different for children and adolescents or for adults, we will focus our analyses on the adult population (≥18 years) [9]. In contrast to Milton and colleagues, we referred to PA in a normal week, as previous studies did [25], rather than in the last week. This reduces the risk of a biased measurement of PA due to circumstances such as recent illness [25]. Furthermore, instead of referring to days per week in which at least 30 minutes of PA is performed, we wanted to refer to the total minutes or total hours per week for a more sensitive measure of total PA.

For analysis we only used the indicated number in minutes per week; reported PA in hours were divided by 60. PA level were categorised in ‘not active’ (0 min/week), ‘low active’ (1-149 min/week), ‘medium active’ (150 – 300 min/week), and ‘very active’ (>300 min/week) [9] according to the recommended WHO guidelines of moderate aerobic PA for the adult population.

To assess smoking status all participants were asked: “Which of the following applies to you best? Please note that smoking means smoking tobacco and not electronic cigarettes or heated tobacco products.” Response options were: (1) I smoke cigarettes every day; (2) I smoke cigarettes but not every day; (3) I do not smoke cigarettes, but I do smoke tobacco in another form (e.g., pipe or cigar); (4) I have stopped smoking completely in the last year; (5) I stopped smoking completely more than a year ago; and (6) I have never been a smoker (i.e., smoked for a year or more). Smoking status were categorised as current smoker = 1-3; ex-smoker = 4-5 and never smoker = 6.

Current smokers were then asked about the number of cigarettes smoked per day. The motivation to stop smoking was measured using the validated German version of the Motivation to Stop Scale [26], a single-item measure that asks: “Which of the following describes you?” with the corresponding response options: (1) I don’t want to stop smoking; (2) I think I should stop smoking but don’t really want to; (3) I want to stop smoking but haven’t thought about when; (4) I really want to stop smoking but I don’t know when I will; (5) I want to stop smoking and hope soon; (6) I really want to stop smoking and intend to in the next 3 months; (7) I really want to stop smoking and intend to in the next month [27]. Current smokers were also asked about their urges to smoke. We first asked: “How much of the time have you felt the urge to smoke in the past 24 hours?” Response options were (1) not at all; (2) a little of the time; (3) some of the time; (4) a lot of the time; (5) almost all the time; (6) all the time [12, 28]. We then asked “ In general, how strong have the urges to smoke been?” with the response options (1) slight; (2) moderate; (3) strong; (4) very strong; (5) extremely strong [12, 28]. Participants who responded 1 (not at all) to the variable time spent with urges to smoke, the variable strength of urges to smoke were coded 0 (i.e., no urges to smoke). Time spent with urges to smoke ranged from 1 (not at all) to 6 (all the time) and strength of urges to smoke from 0 (no urges to smoke) to 5 (extremely strong urges to smoke).

### Statistical analyses

The prevalence of PA level (and corresponding 95% confidence interval, CI) was estimated for the total sample and stratified by age, sex, region of living, educational attainment, income, and smoking status. For these estimations, data were weighted to be representative for the German population. Details on weighting technique have been published in the DEBRA study protocol [29].

To calculate associations between PA levels as the independent variable (not active=reference, low, medium and very active) and smoking outcomes as dependent variables, regression models were applied. For these analyses unweighted data were used, and associations were adjusted for the potential confounding factors age, sex, region, education and income (justification of the chosen factors are published in the study protocol: https://osf.io/cmvk6). We report adjusted odds ratios (OR) and corresponding 95% CI.

To calculate associations between leisure time PA level and smoking status (never smoker=reference, ex- and current smoker) a multinomial logistic regression model was applied. In the subsample of current smokers, associations between leisure time PA level and cigarettes smoked per day were calculated through a multivariable linear regression model. The linear model did not meet the assumption of normal distributed residuals [30]. Therefore, for a more robust result, bootstrapping with 5000 resamples were performed [31]. Additionally, in the subgroup of current smokers, associations between PA level and urges to smoke (strength of urges and times spend with urges in the last 24 hours) as well as the motivation to stop smoking were calculated through three ordinal regression models. In case of missing values, data were deleted and not counted for regression analysis.

## RESULTS

### Sample description

A total of 4,159 individuals aged ≥18 years (unweighted) were interviewed of whom 4,073 (unweighted, 3,921 weighted) answered the question of PA level (97.9%). The prevalence of PA was found to be as follows (Table 1): 29.9% were not active during leisure time, 28.2% low active, 22.1% medium active, 19.7% very active. In the subsample of current smokers, 39.8% were not active during leisure time, 26.7% were low active, 18.1% medium active and 15.3% very active.

**Table 1.** Sample description (total sample) stratified by different physical activity levels (weighted data).

|  | **Total sample<sup>†</sup>** | **Not active**<br>**(0 min/week)** | **Low active**<br>**(1-149 min/week)** | **Medium active**<br>**(150-300 min/week)** | **Very active**<br>**(>300 min/week)** |
| --- | --- | --- | --- | --- | --- |
|  | **100 (3,921)** | **29.9 (1,173)** | **28.2 (1,106)** | **22.1 (868)** | **19.7 (774)** |
| **Sex** |  |  |  |  |  |
| Female | **50.8 (1,993)** | 31.5 (627) | 28.8 (574) | 21.9 (436) | 17.9 (356) |
| Male | **49.2 (1,927)** | 28.3 (545) | 27.6 (532) | 22.4 (432) | 21.7 (418) |
| **Age, year (categories)** |  |  |  |  |  |
| 18-24 | **11.2 (428)** | 18.2 (78) | 24.5 (105) | 28.0 (120) | 29.2 (125) |
| 25-39 | **21.7 (824)** | 21.8 (180) | 31.4 (259) | 26.0 (214) | 20.8 (171) |
| 40-64 | **42.4 (1,612)** | 33.3 (537) | 29.8 (481) | 20.2 (325) | 16.7 (269) |
| 65+ | **24.8 (942)** | 38.2 (360) | 24.3 (229) | 19.3 (182) | 18.2 (171) |
| **Educational attainment<sup>‡</sup>** |  |  |  |  |  |
| Low | **29.2 (1,097)** | 47.7 (523) | 25.0 (274) | 15.4 (169) | 11.9 (131) |
| Middle | **39.4 (1,481)** | 37.2 (468) | 50.8 (441) | 35.2 (339) | 35.2 (233) |
| High | **31.4 (1,178)** | 13.9 (164) | 28.7 (338) | 26.9 (317) | 30.5 (359) |
| **Net monthly household income (€)** |  |  |  |  |  |
| Low | **12.6 (493)** | 42.4 (111) | 22.5 (86) | 17.4 (87) | 17.6 (309) |
| Middle | **62.6 (2,445)** | 31.2 (762) | 30.0 (733) | 21.5 (525) | 17.4 (425) |
| High | **24.8 (969)** | 20.3 (197) | 26.1 (253) | 26.5 (257) | 27.0 (262) |
| **Region of living<sup>~</sup>** |  |  |  |  |  |
| Rural | **40.8 (1,599)** | 30.5 (487) | 30.3 (485) | 21.3 (340) | 17.9 (287) |
| Urban | **43.4 (1,702)** | 29.8 (508) | 27.7 (471) | 24.0 (408) | 18.5 (315) |
| Metropolitan | **15.9 (622)** | 28.8 (179) | 24.1 (150) | 19.3 (120) | 27.8 (173) |
| **Smoking status** |  |  |  |  |  |
| Never | **48.0 (1,878)** | 23.4 (440) | 28.9 (542) | 25.0 (470) | 22.7 (426) |
| Ex | **15.1 (590)** | 26.4 (156) | 29.7 (175) | 23.2 (137) | 20.7 (122) |
| Current | **37.0 (1,447)** | 39.8 (576) | 26.7 (387) | 18.1 (262) | 15.3 (222) |

Data are presented as percentages (absolute numbers). <sup>†</sup>Total sample means all respondents, who answered the question regarding physical activity. Person characteristics stratified by physical activity levels are presented as 100% within the rows. (total sample data are presented as 100% within the columns; sum below or above 100% due to missing values or rounding of cell frequencies). <sup>‡</sup>German equivalents to education attainment listed from lowest to highest: low=no qualification / junior high school equivalent, middle=secondary school equivalent, high=advanced technical college equivalent / high school equivalent. Net monthly household income (OECD equivalent net monthly household income in €). <sup>~</sup>Region of living: rural (<20,000 residents), urban (20,000-500,000 residents), metropolitan (>500,000 residents).

The proportion who were physically inactive was higher among women (31.5%) compared with men (28.3%), older people (38.2% aged over 65 years) compared with younger people (18.2% aged 18-24 years), and people with lower (47.7%) compared with higher educational attainment (13.9%), with lower (42.4%) compared with higher income (20.3%) and people living in metropolitan (28.8%) compared with rural regions (30.5%) (Table 1).

### Associations between leisure time PA level and smoking behaviour

Higher levels of PA were associated with lower odds of being a current smoker, rather than being a never smoker. This was not shown for ex- vs. never smokers.

Among current smokers, higher levels of PA were associated with a lower number of cigarettes smoked per day, weaker strength of urges to smoke, and greater motivation to stop smoking in current smokers. PA level was not significantly associated with the time spent with urges to smoke in the past 24 hours, but only with a small overlap in the upper CI. The results of the associations are shown in Table 2.

**Table 2.**
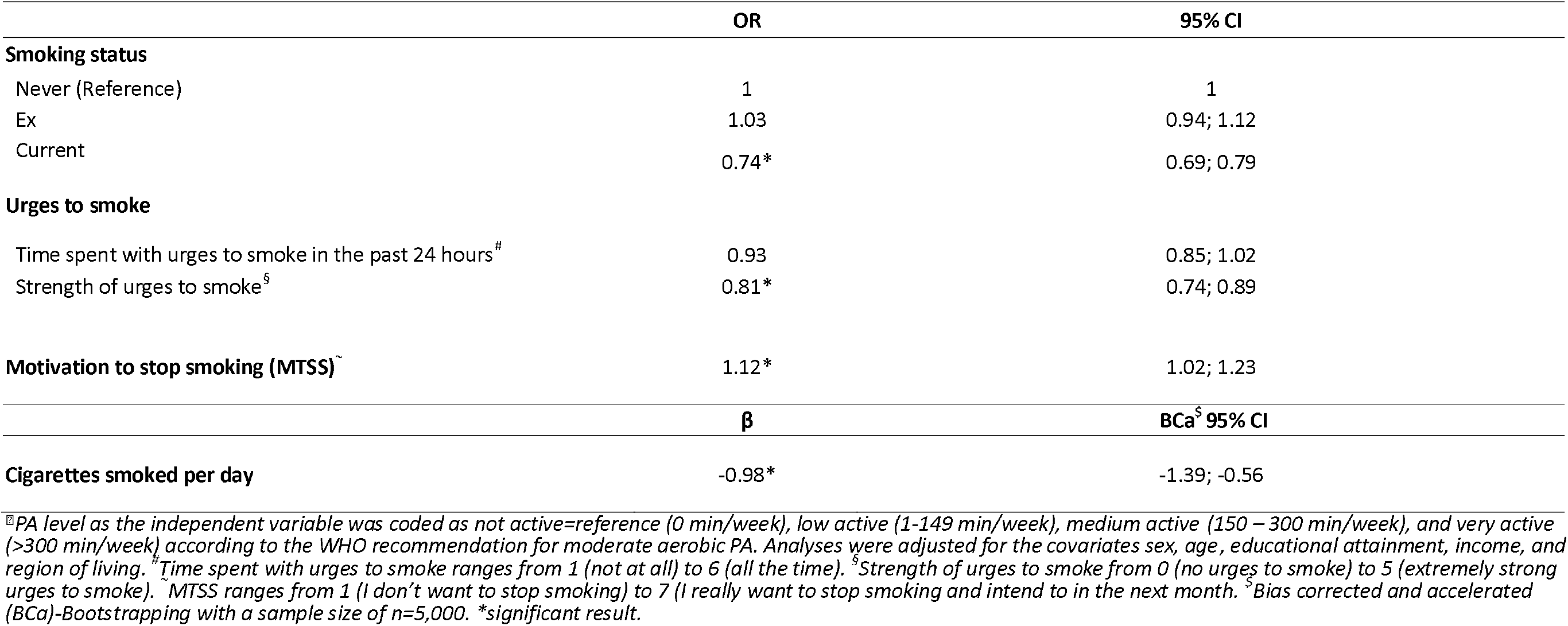
Results of the multinomial logistic regression and ordinal analyses of associations between physical activity levels^⍰^ and smoking behaviour.

## DISCUSSION

Our population survey on smoking behaviour in Germany allowed us to analyse associations between PA levels and smoking behaviour. To complement existing studies that have examined the association between PA levels and smoking status or number of cigarettes smoked per day [3, 10, 11], we additionally considered associations between PA levels and urges to smoke as well as motivation to stop smoking. There was evidence that people with higher PA levels were less likely to be current smokers. Among current smokers, higher PA levels were associated with smoking fewer cigarettes per day, reporting less of an urge to smoke, and being more motivated to stop smoking.

In alignment with results from a study from Switzerland [11], we found that individuals who are physically inactive or engage in low levels of PA during leisure time are more likely to smoke than those who are more physically active. Furthermore, similar to a Norwegian study [11], we showed that current cigarette smokers had lower PA levels compared to never-smokers, despite the difference in the survey regarding the intensity of PA, according to which the authors only included vigorous PA, whereas we combined moderate and vigorous PA.

A dose-response relationship between PA and the number of cigarettes smoked per day has been reported, with higher cigarette consumption associated with an increased risk of low PA [10]. In our study, higher PA levels were also associated with a lower cigarette consumption per day. Our regression analysis yielded a beta value of -0.98 (Table 2), indicating that each one-level increase in PA (e.g., from not to low active, or from medium to very active) was associated with approximately one fewer cigarette smoked per day. A study from China showed a negative effect of tobacco smoking on physiological factors, manifested by a reduction in cardiopulmonary endurance through a decrease in maximal oxygen uptake (VO_2_max) in young college students aged 19-26 years [32]. As cigarette smoking has an immediate negative effect on cardiovascular function during exercise [33], it is possible that individuals who engage in aerobic exercise are more likely to be non-smokers or to smoke fewer cigarettes per day than individuals who do no or less aerobic exercise or even strength training. Whether there is an association between different PA intensities, durations or types of PA and the number of cigarettes smoked per day should be investigated in future studies.

Consistent with the results of previous studies investigating the effect of PA interventions on nicotine craving, higher PA levels were associated with weaker urges to smoke among smokers in our study [15, 17]. We also found that higher PA levels was associated with lower strength of urges to smoke, but not statistically significant with the time spent with urges to smoke in the last 24 hours. There was some overlap in the upper CI (Table 2), making the result inconclusive. The point estimate was in the same direction as the other variables, but was small, so it could just be a small association and we did not have the sample size to detect it. An additional question about PA in the last 24 hours which would address the same time period could may yield different outcomes.

Furthermore, in line with an interventional study [14], our data showed that higher PA levels are associated with greater motivation to stop smoking. This could be due to various reasons, which can be both physiological (e.g. lung function) and psychological (e.g. changes in mood) [15]. For example, physiologically, an increase in PA may be perceived by current smokers as having a negative effect on the smoking-induced decline in lung function [32], thus increasing motivation to quit smoking. On the other hand, people who have successfully quit smoking may be more likely to adopt a healthier lifestyle, including increasing their PA [34]. The extent to which physiological or psychological factors are associated with the motivation to stop smoking at different levels and types of PA remains uncertain and may vary between individuals. However, we have been able to show through our population based study that people with a generally higher PA level seem to be more motivated to stop smoking, rather than people with lower PA level. The extent to which PA alone or other reasons contribute to increased motivation to stop smoking should be investigated in further studies. Knowing the impact of PA on the motivation to stop smoking can help in designing PA interventions or smoking cessation programmes.

### Strengths and limitations of the study

Our study design with cross-sectional data does not allow for causal inferences. In addition, the single-item question does not allow for differences in the intensity, duration, frequency, and type of PA (e.g. strength or endurance training). Therefore, people who met the WHO recommendation based on vigorous PA (75 min/week) for aerobic exercise, were classified here as not meeting the recommendation for moderate PA (150 min/week) and may have been misclassified as low active. Thus, multiple items should be included to distinguish between moderate and vigorous aerobic PA, and the type of PA as these differences may influence smoking behaviour. We also do not know the specific reasons for quitting smoking or increasing PA. The reasons for adopting a healthier lifestyle vary from person to person and are related to different personal circumstances. Key strengths of the study include the nationally representative population sample and the breadth of data on smoking behaviour.

### Research/ Policy implications

Understanding how different PA behaviours are associated with smoking behaviour can help develop prevention strategies and inform specific target specific groups such as current smokers. In addition, knowledge of these associations may help in the design of public health campaigns. Further research is needed to show whether differences in PA behaviours (e.g. intensity, duration and frequency) lead to differences in smoking behaviour, and whether this is also reflected when comparing PA in leisure time and at work.

## CONCLUSION

PA levels are associated with different smoking behaviours. People who are more physically active are less likely to smoke. Current smokers who are more physically active smoke less, are less dependent, and are more motivated to quit. Further research is needed to investigate whether the observed associations are causal and whether changes in PA lead to changes in smoking outcomes.

## Funding

The DEBRA study was funded from 2016-2019 (waves 1-18) by the Ministry of Innovation, Science and Research (MIWF) of the State of North Rhine-Westphalia as part of the “NRW Return Program”. Since 2019 (from wave 19), the study has been funded by the Federal Ministry of Health.

## Competing interests

None declared.

## Patient consent for publication

Not applicable.

## Ethics approval

The DEBRA study has been approved by the Ethics Committee at the Heinrich-Heine-University Duesseldorf, Germany (ID 5386/R), and has been registered at the German Clinical Trials Register (<u>DRKS00017157</u>).

## Data availability statement

Data are available upon reasonable request. The data supporting this study will be made available upon reasonable request to the corresponding author.

## Notes

### Competing Interest Statement

The authors have declared no competing interest.

### Clinical Protocols

https://osf.io/7gzwd/

### Funding Statement

The study was funded by the Federal Ministry of Health.

### Author Declarations

Ethics Committee at the Heinrich-Heine-University Duesseldorf, Germany gave ethical approval for this work (ID 5386/R)

